# Atypical Resting State Functional Connectivity in Mild Traumatic Brain Injury

**DOI:** 10.1101/2021.04.26.21256114

**Authors:** Joelle Amir, Jay Kumar Raghavan Nair, Raquel Del Carpio, Alain Ptito, Jen-Kai Chen, Jeffrey Chankowsky, Simon Tinawi, Ekaterina Lunkova, Rajeet Singh Saluja

## Abstract

**Objectives:** This study aimed to investigate changes in three intrinsic functional connectivity networks (IFCNs; default mode (DMN), salience (SN), and task-positive networks (TPN)) in individuals who had sustained a mild traumatic brain injury (mTBI).

**Methods:** Resting-state functional magnetic resonance imaging (rs-fMRI) data was acquired from 27 mTBI patients with persistent post-concussive symptoms (PCS), along with 26 age- and sex-matched controls. These individuals were recruited from a Level-1 trauma centre, at least three months after a traumatic episode. IFCNs were established based on seed-to-voxel, region-of-interest (ROI) to ROI, and independent component analyses (ICA). Subsequently, we analyzed the relationship between functional connectivity and PCS.

**Results:** Seed-to-voxel analysis of rs-fMRI demonstrated decreased functional connectivity in the right lateral parietal lobe, part of the DMN, and increased functional connectivity in the supramarginal gyrus, part of the SN. Our TPN showed both hypo- and hyperconnectivity, dependent on seed location. Within network hypoconnectivity was observed in the visual network, also using group comparison. Using an ICA, we identified altered network functional connectivity in regions within four IFCNs (sensorimotor, visual, DMN, and dorsal attentional). A significant negative correlation between dorsal attentional network connectivity and behavioural symptoms score was also found.

**Conclusions:** Our findings indicate that rs-fMRI may be of use clinically, in order to assess disrupted functional connectivity among IFCNs in mTBI patients. Improved mTBI diagnostic and prognostic information could be especially relevant for athletes looking to safely return to play, as well for individuals from the general population with persistent post-concussive symptoms months after injury, who hope to resume activity.

## 1. Introduction

In recent years, mild traumatic brain injury (mTBI) has emerged as a significant global public health concern. Although mTBI lies at the mildest end of the TBI spectrum, it is a misnomer as it accounts for more than one million emergency department visits annually in the United States alone (D’Souza et al., 2020; Iraji et al., 2015). There is therefore a substantial burden faced not only by the affected individuals and their loved ones, but also by the healthcare system. To further complicate matters, conventional neuroimaging results are often normal in mTBI, posing a constant challenge for physicians to provide prognostic information (Iraji et al., 2015; Wu et al., 2016). MTBI has therefore frequently been labeled a “silent epidemic” because, despite negative findings, mTBI patients can develop long-term symptoms that do not resolve by three months post-injury (Yan et al., 2017). The constellation of symptoms have been termed persistent post-concussive symptoms, characterized by physical, cognitive, and behavioural disturbances that may take up to a year to return to baseline, if at all (Bharath et al., 2015). The pathophysiology of post-concussive symptoms is still poorly understood, and there is therefore an urgent need for new, more objective tools that may complement the data provided by Computed Tomography and Magnetic Resonance Imaging (MRI) as the standard evaluation techniques in mTBI (Wu et al., 2016; Zhou et al., 2012).

In the past few decades, the number of studies using advanced neuroimaging techniques to investigate mTBI has grown exponentially (Wu et al., 2016). In particular, resting state functional MRI (rs-fMRI) has allowed for the evaluation of intrinsic functional connectivity networks (IFCNs) which represent the maintenance of baseline energy expenditure in the brain (Zhou et al., 2012). The default mode network (DMN) is the most frequently cited among all IFCNs, and it is often studied in conjunction with the executive or task-positive network (TPN), which demonstrates an anti-correlated relationship at rest (Iraji et al., 2015, Mayer et al., 2011). A third network known as the salience network (SN) has been shown to mediate the balance between DMN and the executive network, which may be disrupted following mTBI (Sours et al., 2013, Sharp et al., 2014). Although functional connectivity changes in these networks may prove useful as biomarkers of brain injury, limited cohesiveness among data has been reported (Chong & Schwedt, 2018). Inhomogeneities such as TBI history, time since injury, and analysis methods are just some of the confounding factors making it difficult to compare findings (Johnson et al., 2012, Puig et al., 2020). Importantly, the majority of the previous literature has focused on a specific subset of IFCNs, without investigating what other, perhaps less common, networks may be involved in the pathogenesis of mTBI. It is therefore imperative to include complementary analysis approaches that taken together may provide insight into between network connectivity, within network connectivity, and whole-brain connectivity disruptions underlying mTBI.

The current study aims were to a) to examine alterations in functional connectivity between and within two IFCNs (DMN and SN), the TPN, and other brain regions using a seed-based analysis sample of mTBI patients compared to matched healthy controls; and b) to investigate what other IFCNs, if any, are implicated in mTBI, using an independent component analysis (ICA). We expect to find network-dependent functional connectivity changes to various degrees in all three IFCNs, as well as in other visually identifiable IFCNs. IFCNs may present with either functional connectivity deficits, excesses pointing to increased recruitment of neural resources due to injury, or both. A supplemental aim will be to assess the relationship between functional connectivity and severity and type of post-concussive symptoms. In using multiple methods of analysis, this approach has the potential to detect subtle changes which underlie the symptoms associated with mTBI, thereby supporting the use of disrupted functional connectivity among IFCNs as a promising outcome predictor following injury.

## 2. Materials & Methods

### 2.1 Participants

A cohort of 27 mTBI patients aged 18-60 years was referred by the Mild Traumatic Brain Injury Clinic at the Montreal General Hospital, a Level-1 trauma centre. They had persistent cognitive, emotional, and functional disturbances even after symptomatic treatment, and were recruited at three months after the traumatic episode. For comparison, 26 age- and sex-matched controls were also enrolled. Patient eligibility was based on the mTBI definition by the World Health Organization Collaborating Centre for Neurotrauma Task Force as well as a Rivermead Post Concussion Symptoms Questionnaire (RPQ) score greater than 30. Patients and healthy controls were excluded if they had a) a head injury within the past year or were continually suffering symptoms from a previous head injury at the time of injury; b) previous history of neurological disorders; c) substance abuse problems; or d) contraindications for MRI (i.e. claustrophobia, metallic implants, etc.). Approval from the McGill University Health Centre ethics review board was obtained before commencement of the study, and patients signed informed consent forms prior to participation. Data will be made openly available from the corresponding author upon reasonable request.

**Table 1:** Demographics of mTBI and control subjects.

|  | mTBI (n = 27) | Control (n = 26) |
| --- | --- | --- |
|  | Mean (SD), range |  |
| **Age (at MRI, y)** | 43.9 (10.2), 28-60 | 43.2 (10.2), 23-60 |
| **Time since injury (mo)** | 8.8 (6.0), 3.2-21.4 | n/a |
| **Rivermead score** | 31.3 (12.2) | n/a |
| Physical symptoms | 17.4 (6.1) |  |
| Cognitive symptoms | 7.3 (3.3) |  |
| Behavioural symptoms | 6.3 (4.7) |  |
|  | n (%) |  |
| **Gender** |  |  |
| Male | 8 (30) | 8 (31) |
| Female | 19 (70) | 18 (69) |
| **Mechanism of Injury** |  | n/a |
| Motor vehicle collision | 7 (26) |  |
| Pedestrian/cyclist vs. car | 4 (15) |  |
| Fall from own height | 3 (11) |  |
| Fall from height (> 2m) | 5 (19) |  |
| Fall down stairs | 2 (7) |  |
| Hit head against object | 4 (15) |  |
| Assault | 2 (7) |  |
| **Number of Previous mTBIs** |  | n/a |
| 0 | 20 (74) |  |
| 1 | 5 (19) |  |
| 2 | 0 |  |
| 3 | 2 (7) |  |

### 2.2 Cognitive assessments

Cognitive assessments were carried out on all subjects, using the Standardized Assessment of Concussion. This tool was developed in order to obtain a more objective assessment of an athlete’s cognitive state on the sidelines immediately following concussion, but has been deemed useful in the general population as well (McCrea, 2001). It includes evaluations of attention, orientation and memory (Yan et al., 2017). The presence and severity of post-concussive symptoms were assessed by the RPQ. The questionnaire is comprised of 16 symptoms that frequently occur after brain injury. Subjects were asked to rate the severity of these symptoms over the last 24 hours from 0 (no or no more symptoms than before the trauma) to 4 (symptoms of highest severity). The sum of the scores for the 16 symptoms were then obtained. Symptoms were also grouped into three sub-categories, which includes physical (e.g., headaches, dizziness, nausea, vomiting, noise sensitivity, sleep disturbance, fatigue, blurred vision, light sensitivity, double vision), cognitive (e.g., poor concentration, poor memory, increased thinking time), and behavioural symptoms (e.g., irritability, depression, frustration, restlessness) each with a total score of 36, 12, and 16 respectively.

### 2.3 Image acquisition

All subjects underwent MRI examinations at the Royal Victoria Hospital. Rs-fMRI data was acquired on a 3-Tesla Siemens Skyra scanner with a 32-channel radiofrequency head-only coil using gradient echo-planar T2*-weighted imaging sequence (time repetition = 2250 ms, time echo = 30 ms, flip angle = 90°, matrix size = 64 x 64, FOV = 225 mm), with 42 oblique slices covering the whole brain (slice thickness = 3.5mm isotropic). For anatomical reference, a high-resolution T1-weighted image was also acquired for each subject (3D MP-RAGE, TR = 2300ms, TE = 2.98 ms, 176 slices, slice thickness = 1mm, FOV = 256mm, image matrix = 256 x 256, flip angle = 9 degrees, interleaved excitation). The resting state scan duration was eight minutes, during which 210 fMRI volumes were acquired. The subjects were instructed to keep their eyes closed and stay awake; after every sequence, the technologist would ask the patients how they were feeling, which helped ensure that they did not fall asleep.

### 2.4 Pre-processing

MRI data was preprocessed using SPM 12 within the Functional Connectivity (CONN) toolbox (http://www.nitric.org/projects/conn, version 19.c), using the “direct normalization to MNI space” pipeline. Data were functionally realigned and unwrapped; slice-time corrected; structurally and functionally normalized and segmented into grey matter, white matter and CSF tissue; flagged as potential outliers if framewise displacement exceeded 0.9 mm or if global BOLD signal changes were above five standard deviations; and smoothed, using an 8mm Gaussian kernel (Whitfield-Gabrieli & Nieto-Castanon, 2012). Both functional and structural data were resampled with 2 mm and 1 mm voxels, respectively. Artifacts were detected using the Artifact Removal Tools toolbox and entered into a linear regression as potential confounding effects (white matter, CSF, realignment, scrubbing, and effect of rest) in order remove any influence on the BOLD signal. After linear de-trending was performed, images were then band-pass filtered to 0.008Hz – 0.09Hz and motion regressed to minimize the effect of motion and noise sources. No significant group differences were found for average realignment (p = 0.6544) or framewise displacement (p = 0.5278).

### 2.5 Seed-based analysis

A seed-based correlation approach was used to evaluate the BOLD signal associated with functional connectivity of the DMN, the TPN, and the SN. Based on previous studies, the seeds for functional analysis were placed within the medial prefrontal cortex (MPFC), the left and right lateral parietal lobes, and the posterior cingulate cortex (PCC) to determine connectivity within the DMN, whereas the left and right dorsolateral prefrontal cortex (DLPFC), the left and right insula, the supplementary motor area (SMA), the left and right inferior parietal lobes, the left and right thalamus, were used as seeds to define the TPN. The TPN nodes were selected based on unpublished fMRI data by our group on 100 healthy McGill athletes while performing a working memory task (see Chen et al. 2004 for details of the task). Finally, the SN seeds were placed in the anterior cingulate cortex (ACC), the left and right anterior insula, the left and right rostral prefrontal cortex, and the left and right supramarginal gyri. The selected DMN and SN IFCN nodes are part of CONN’s default resting state nodes, which include 32 seeds/targets. The size of the seeds used correspond to one voxel (1 x 1 x 1 mm^3^).

The CONN toolbox performs seed-based analysis by comparing the temporal correlation of the extracted BOLD signal to all remaining voxels in the brain, as well as ROI-based analysis by grouping voxels into ROIs based on Brodmann areas (Johnson et al., 2012). Therefore, all Brodmann areas were imported as potential connections for the chosen seed ROIs. To manually create the TPN ROIs, we entered the MNI coordinates for each TPN seed, using a spherical radius of 6 mm. In order to validate multiple comparisons, Fisher transformed *z* scores were used along with SPM functions within the CONN toolbox. These ROI-based analyses were performed on all subjects with a general linear model test to determine significant DMN, TPN, and SN connections at the individual level (1^st^-level analysis). Based on the results obtained in the 1^st^-level analysis, an unpaired t-test was performed with a threshold set at p<0.05 false discovery rate (FDR) corrected, as well as a voxel-wise p<0.001 (uncorrected), to determine significantly different connections between healthy controls and mTBI groups (2^nd^-level analysis). Finally, within network ROI-to-ROI connectivity was also performed, by calculating the average pairwise connectivity measures for all eight default networks in CONN.

### 2.6 Group ICA

A group ICA was run with a FastICA for estimation of independent spatial components and GICA1 back-projection for spatial map estimation at the individual subject level. Dimensionality reduction was set to 64 and number of components was set to 40 as per CONN’s default settings. A technique known as the correlational spatial match-to-template approach was employed within CONN to identify each component of the brain IFCNs for the default mode, sensorimotor, visual, salience, dorsal attention, frontoparietal, language and cerebellar networks. Group ICA uses a dual regression approach, through the unification of three stages. First, ICA decomposes the data by recognizing distinct patterns of functional connectivity in each subject. Next, spatial maps and associated time courses are identified for each subject. Finally, component maps are generated and are compiled in order to perform the non-parametric analysis, which allows statistical significance to be extracted across groups (Smitha et al., 2017). Significant group differences within each identified component can then be determined (2^nd^-level analysis).

### 2.7 Correlation with post-concussive symptoms

Additional statistical analyses were performed in SPSS 26. To determine correlations with RPQ scores, as well as physical, cognitive, and behavioural symptoms, we extracted ROIs where significant group differences were found between mTBI patients and healthy controls in the seed-based and group ICA analyses, using rex, a tool within CONN. These values were then entered into SPSS along with RPQ scores and the three sub-categories (physical, cognitive, behavioural), and a linear regression was performed. Age and sex were also included as confounds.

### 2.8 Effect of injury history

Injury history was controlled for in all analyses by entering the number of previous TBIs as a covariate in an ANCOVA, along with the extracted ROIs where significant group differences were found in the seed-based and group ICA analyses.

## 3. Results

### 3.1 Seed-to-voxel results

The DMN in mTBI demonstrated an overall reduced connectivity compared to controls (p-FDR corrected < 0.05, p-uncorrected < 0.001). For the right lateral parietal lobe seed, the BOLD connectivity was greater for controls compared to mTBI patients specifically within the precuneus. No significantly different connections were found for the MPFC or the PCC seed for group comparison.

For the TPN, results varied across seeds. For the right DLPFC seed, the BOLD connectivity was greater in mTBI patients compared to controls, specifically within the right lateral occipital cortex. For the right insula, the BOLD connectivity was greater in controls compared to mTBI patients, particularly within the left and right lateral occipital cortex, as well as within the cingulate gyrus. No significantly different connections were found for the remaining seven TPN seeds for group comparison.

For the SN, the BOLD connectivity was greater in mTBI patients compared to controls, for the right supramarginal gyrus, specifically within the right lateral occipital cortex and the right superior parietal lobule. No significantly different connections were found for the remaining six seeds for group comparison.

**Table 2:** Seed-to-voxel results showing differences in functional connectivity between mTBI patients and healthy controls, after controlling for the effect of injury history (uncorrected threshold p < 0.01, FDR-corrected threshold p < 0.05).

| Seed | Target(s) | Voxels | Peak MNI coordinates | Peak *t* statistic | *p* (FDR) | Direction of FC change relative to controls |
| --- | --- | --- | --- | --- | --- | --- |
| DMN |  |  |  |  |  |  |
| Lateral parietal R | Precuneus | 226 | 10 -52 74 | -4.77 | 0.000014 | Hypoconnectivity |
| TPN |  |  |  |  |  |  |
| DLPFC R | Lateral occipital cortex R | 66 | 52 -62 -6 | 3.95 | 0.043118 | Hyperconnectivity |
| Insula R | Lateral occipital cortex R | 92 | -34 -86 -16 | -5.28 | 0.015997 | Hypoconnectivity |
|  | Lateral occipital cortex L | 25 | 38 -78 -10 |  | 0.023159 |  |
|  | Cingulate gyrus | 58 | -4 -10 46 |  | 0.045050 |  |
| SN |  |  |  |  |  |  |
| Supramarginal gyrus R | Lateral occipital cortex R | 69 | 40 -78 16 | 4.49 | 0.013429 | Hyperconnectivity |
|  | Superior parietal lobule R | 110 | 24 -54 56 |  | 0.013429 |  |

**Figure 1:**
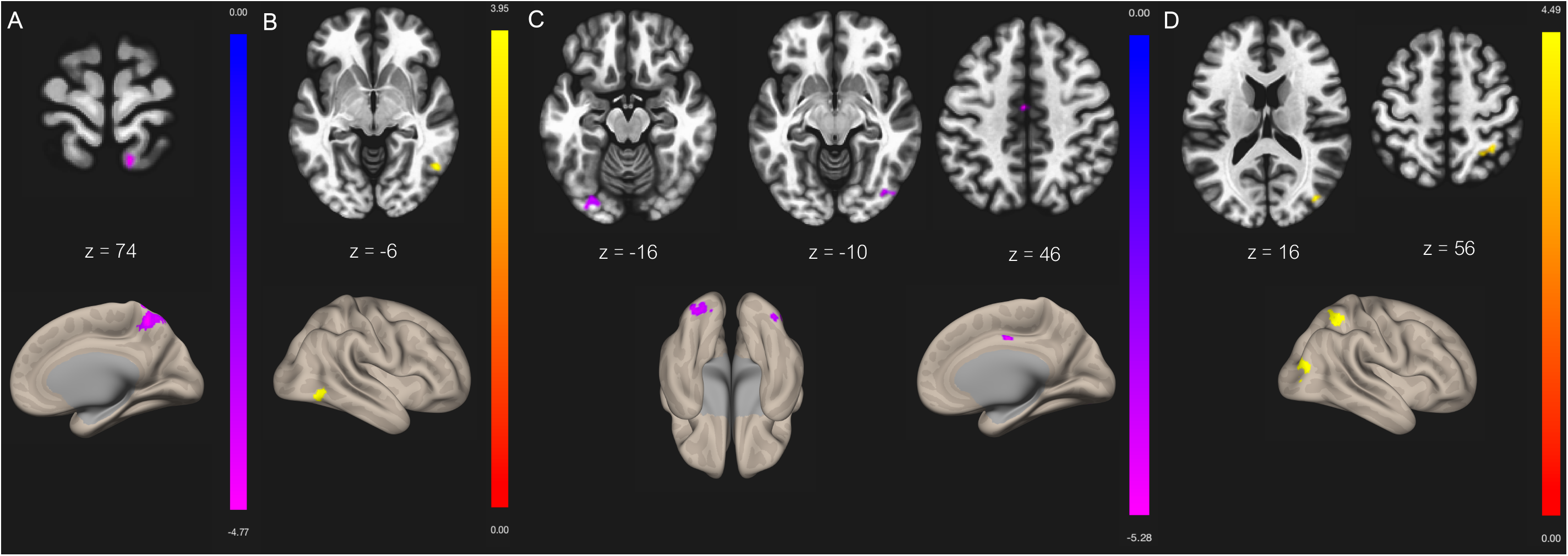
Seed-to-voxel contrast between mTBI > healthy controls FC for **A)** lateral parietal (R) with precuneus, **B)** DLPFC (R) with lateral occipital cortex (R), **C)** insula (R) with lateral occipital cortex (R), lateral occipital cortex (L), and cingulate gyrus, **D)** supramarginal gyrus (R) with lateral occipital cortex (R) and superior parietal lobule (R). Slices denote the peak activation coordinates where the color bar represents positive t-values in yellow/red and negative t-values in blue/purple. Slices are displayed at uncorrected threshold p < 0.001, and FDR-corrected cluster threshold p < 0.05.

### 3.2 ROI-to-ROI results

No ROI-to-ROI connectivity differences were found between controls and mTBI patients when using FDR correction for the DMN, the TPN or the SN. Further analyses were run on the remaining five default networks in CONN, and group differences were found only within the visual network. In the right lateral seed of the visual network, mTBI patients demonstrated hypoconnectivity compared to controls, with decreased connections to both the left lateral seed and the medial seed of the visual network (p-FDR corrected = 0.042535).

**Figure 2:**
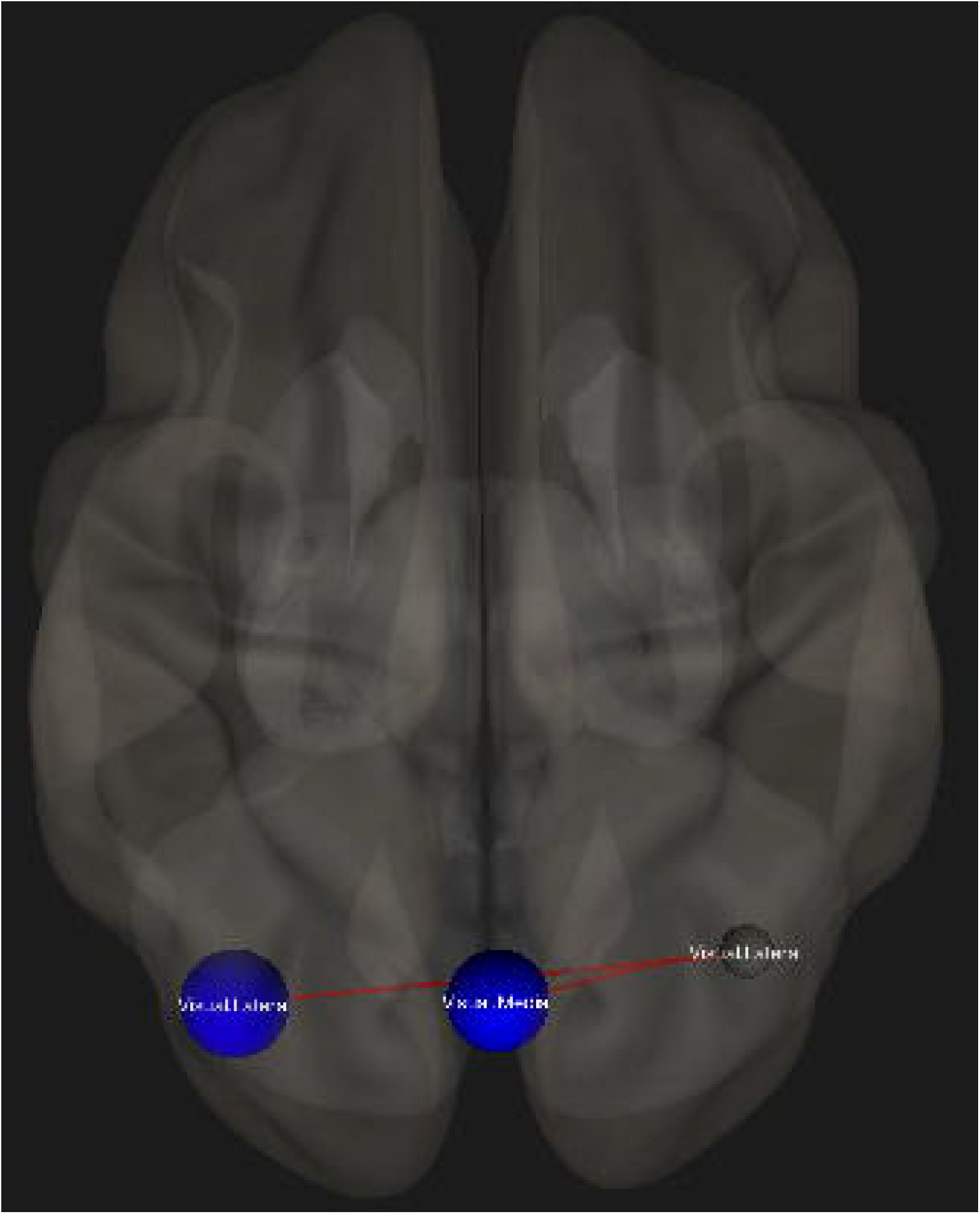
ROI-to-ROI contrast showing decreased FC (blue) for the mTBI group as compared to healthy controls, after controlling for the effect of injury history. The graphic is displayed in superior view with ROI-to-ROI connection threshold set at p-FDR corrected < 0.05.

### 3.3 ICA results

Results from the ICA showed abnormal mTBI functional connectivity in regions within four out of eight brain networks identified by CONN (p-FDR corrected < 0.05, p-uncorrected < 0.001). Decreased functional connectivity was found in mTBI patients compared to controls between the visual network and the right temporal occipital fusiform cortex. The visual network also showed increased functional connectivity with the right lateral occipital cortex in mTBI patients compared to healthy subjects. Increased functional connectivity was additionally found between the following areas: the sensorimotor network and the left frontal orbital cortex, the DMN and the precuneus and left cuneal cortex, and the dorsal attention network and the cingulate gyrus.

**Table 3:**
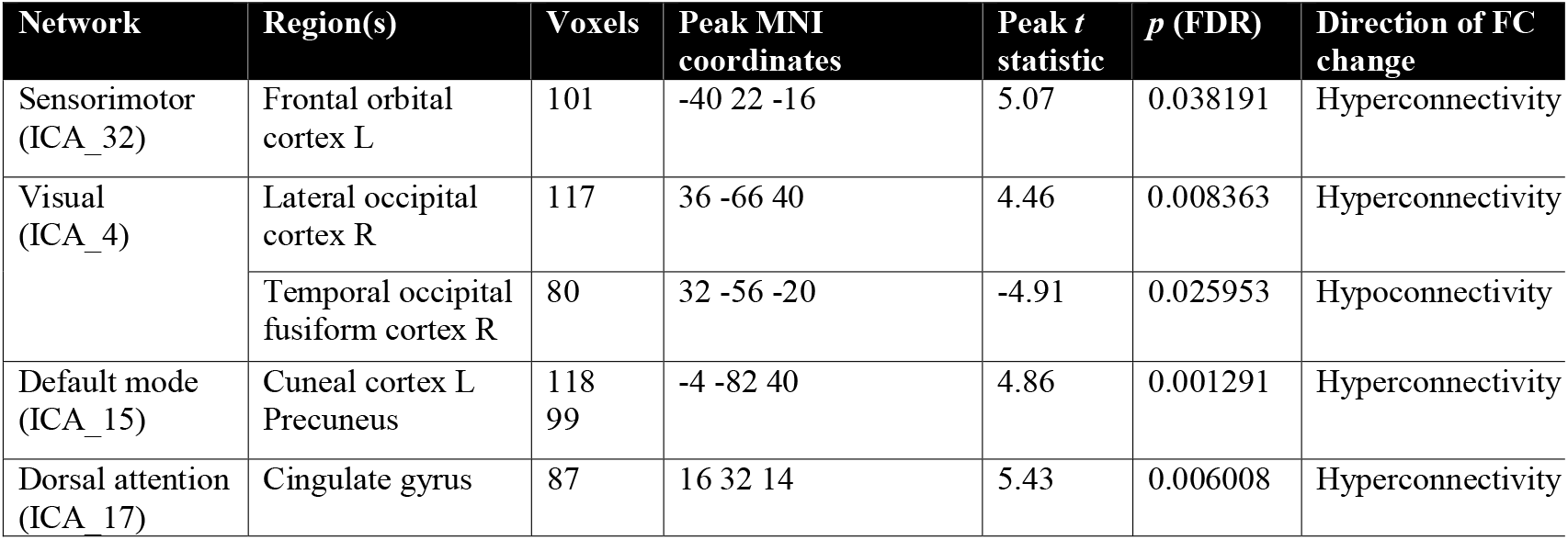
Group ICA results showing differences in functional connectivity between mTBI patients and healthy controls, after controlling for the effect of injury history (uncorrected threshold p < 0.01, FDR-corrected threshold p < 0.05).

| Network | Region(s) | Voxels | Peak MNI coordinates | Peak *t* statistic | *p* (FDR) | Direction of FC change |
| --- | --- | --- | --- | --- | --- | --- |
| Sensorimotor (ICA_32) | Frontal orbital cortex L | 101 | -40 22 -16 | 5.07 | 0.038191 | Hyperconnectivity |
| Visual (ICA_4) | Lateral occipital cortex R | 117 | 36 -66 40 | 4.46 | 0.008363 | Hyperconnectivity |
|  | Temporal occipital fusiform cortex R | 80 | 32 -56 -20 | -4.91 | 0.025953 | Hypoconnectivity |
| Default mode (ICA_15) | Cuneal cortex L<br>Precuneus | 118<br>99 | -4 -82 40 | 4.86 | 0.001291 | Hyperconnectivity |
| Dorsal attention (ICA_17) | Cingulate gyrus | 87 | 16 32 14 | 5.43 | 0.006008 | Hyperconnectivity |

**Figure 3:**
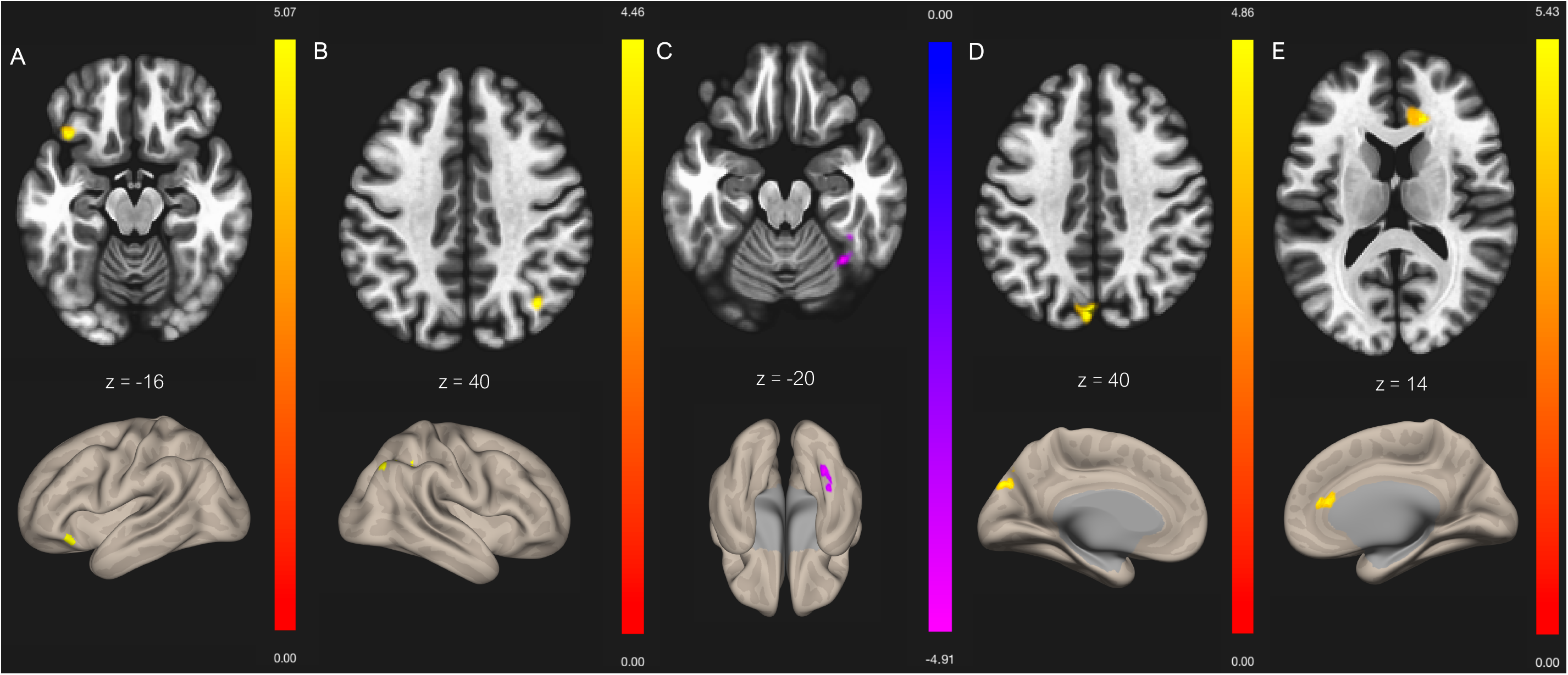
ICA contrast between mTBI > healthy controls FC for **A)** sensorimotor network with frontal orbital cortex (L), **B)** visual network with lateral occipital cortex (R), **C)** visual network with temporal occipital fusiform cortex (R), **D)** DMN with cuneal cortex (L), **E)** dorsal attentional network with cingulate gyrus. Slices denote the peak activation coordinates where the color bar represents positive t-values in yellow/red and negative t-values in blue/purple. Slices are displayed at uncorrected threshold p < 0.001, and FDR-corrected cluster threshold p < 0.05.

### 3.4 Correlation with post-concussive symptoms

Nine regions were tested against RPQ scores, based on the networks that showed significant differences between groups in the analyses mentioned above. After age and sex were taken as confounds, a significant negative relationship was found between the dorsal attentional network and behavioural symptoms score (p = 0.05). No other significant correlations were found between any network and RPQ score or sub-categories.

**Figure 4:**
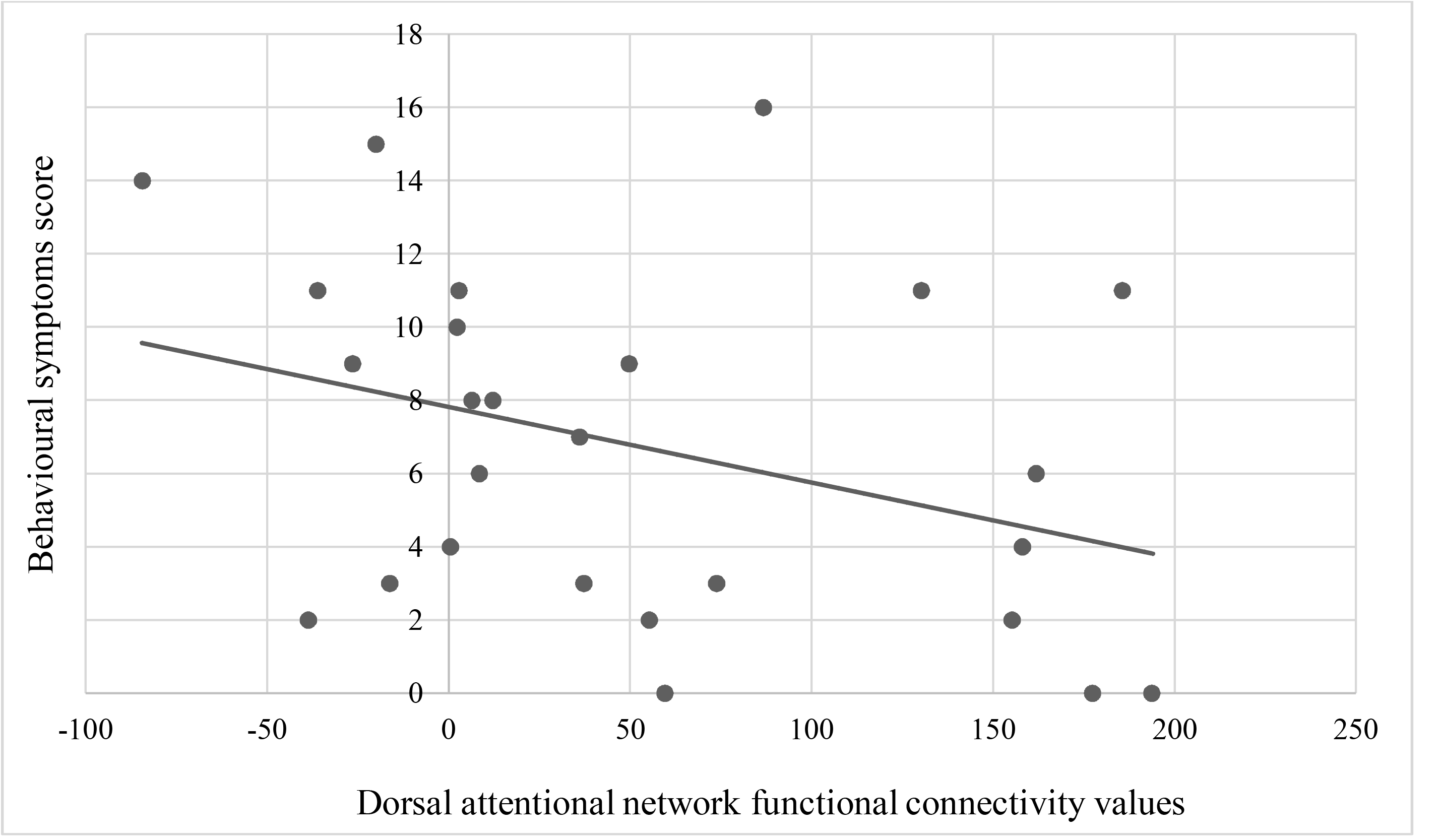
Significant negative correlation between the dorsal attentional network functional connectivity values and behavioural symptoms score (p = 0.05), as demonstrated by the trendline.

## 4. Discussion

We investigated functional connectivity changes in mTBI patients with persistent post-concussive disturbances at least three months after injury, using seed-based, ROI-to-ROI and ICAs. There were four major findings. First, the seed-to-voxel analysis displayed decreased functional connectivity within two DMN nodes, whereas the SN demonstrated increased functional connectivity. Our TPN showed both increased and decreased functional connectivity, dependent on TPN seed origin. Second, the ROI-to-ROI connectivity analysis demonstrated within network connectivity in visual network nodes. Third, using ICA, we identified altered network functional connectivity in regions within five IFCNs: the language network, the sensorimotor network, the visual network, the DMN, and the dorsal attentional network. Finally, the dorsal attentional network was the only network that correlated with post-concussive symptoms score.

### 4.1 Seed-to-voxel findings

One of the most straightforward techniques being implemented for analysing rs-fMRI data is to extract the BOLD signal from a given ROI, in order to compute the temporal correlation between this specific voxel and all other voxels in the brain (Storti et al., 2013). Our seed-to-voxel analysis using this method revealed several findings. Firstly, hypoconnectivity in the DMN is consistent with previous studies, that also found connections from DMN nodes to the left supramarginal gyrus, among others (Sours, Zhuo, et al., 2015). The DMN has been linked to the integration of cognitive and emotional processing, environmental monitoring, and day dreaming, all which may be disrupted following mTBI (van den Heuvel & Hulshoff Pol, 2010). Only the right lateral parietal lobe demonstrated altered functional connectivity in our analysis after controlling for injury history. This region has been considered essential in a number of functions, such as movement planning and control, multisensory integration, and even episodic memory retrieval, which may also be disturbed post-injury (Johnson et al., 2012). Although we did not find significant differences for the MPFC or the PCC similar to other reports, the general consensus across studies points toward reduced functional connectivity within the DMN as a reflection of axonal disruption to white matter pathways connecting various nodes of this network (Sours, George, et al., 2015).

For the DLPFC seed of our TPN, BOLD connectivity in mTBI patients exceeded that of healthy subjects. The level of functional connectivity of the DLPFC has been found to be a powerful predictor for cognitive performance (Song et al., 2008), and has been shown to be proportional to symptom severity in concussed individuals, through task-based experiments by our group (Chen et al., 2007, Chen et al., 2008). Several studies (Johnson et al., 2012; Sours et al., 2013; Sours et al., 2015) found increased functional connectivity between the DLPFC and nodes of the DMN in the mTBI population, whereas our DLPFC seed specifically targeted the lateral occipital cortex. Inhomogeneities are likely because we manually created the ROIs of our TPN seeds, based on existing data from healthy athletes. Nonetheless, this hyperactivity could represent a compensation mechanism due to an increased need for the TPN to have top-down control in suppressing the DMN, which may be necessary in preventing internally-focused, potentially disruptive, thoughts (Sours et al., 2013). The second seed in our TPN that demonstrated significant findings was the right insula. Previous reports have shown functional connectivity abnormalities in this region, such as decreased volume and cerebral blood flow, with changes related to cognitive assessment scores (Lu et al., 2020). Although no correlation was found between the insula and RPQ scores in our study, alterations in this region may nonetheless help pinpoint cognitive dysfunction following mTBI.

The DMN has been shown to correlate negatively with the SN (Di & Biswal, 2015), which coincides with our results, as BOLD connectivity was greater in mTBI patients in the SN compared to controls. The SN is known to respond to external events that are behaviourally salient, and in doing so, appears to subsequently reduce excessive DMN activity (Sharp et al., 2014). The dysfunction of the SN will disrupt the activity of other networks and, therefore, proper functioning is necessary for the control of cognitive processes (Smitha et al., 2017).

Mayer et al. (2011) suggests that reduced connectivity within the DMN and increased connectivity in regions associated with top-down control, such as the SN, “may provide a physiological substrate for common but poorly understood neuropsychiatric complaints following mTBI” (p. 7). It is therefore not surprising that the DMN demonstrated reduced functional connectivity, while nodes of our TPN and SN showed increased functional connectivity in mTBI subjects.

### 4.2 ROI-to-ROI findings

ROI-to-ROI functional connectivity differences were found within the visual network, specifically from the right lateral seed to the left lateral seed and to the medial seed. Even though reduced interhemispheric connectivity within the visual network in mTBI subjects is consistent with previous reports (Li et al., 2020; Slobounov et al., 2011), we may still question why a difference in functional connectivity exists solely within this network. Many areas within the visual cortex are in fact vulnerable to mTBI, as almost 70% of sensory processing is related to vision (Capó-Aponte et al., 2017). Despite the fact that these impairments are most often observed in the acute phase (Slobounov et al. 2011), Gilmore et al. (2016) found decreased visual connectivity one to five years after blast-induced mTBI; this shows that deficits may still be possible for extended periods after injury. Although it is not uncommon for the visual network to be overlooked in mTBI assessment, dysfunction in visual integration following concussion is especially concerning for athletes, as proper functioning is necessary in order to avoid re-injury (Churchill et al., 2017).

### 4.3 ICA findings

While the seed-to-voxel analysis calculates only the functional connectivity between network nodes, the ICA performs a voxel-to-voxel level analysis assessing the connectivity of the brain as a whole (Slobounov et al., 2011). ICA is an attractive technique as it can easily identify functional connections within regions that are not restricted to the boundaries of these nodes (Kornelsen et al., 2020). This method therefore has the ability to capture an entire network as a single major component, and can separate whole-brain signal fluctuations from physiological noise (De Luca et al., 2006). In turn, this can decrease the heterogeneity of patterns within a network that may occur when using a seed-based technique (Greicius et al., 2003).

Decreased functional connectivity was found in one network using the ICA: the visual network. A substantial amount of stored information in the human brain is presumed to be related to visual processes, as this network serves to integrate sensory information that is relayed to higher order executive functions (Gilmore et al., 2016). In fact, decreased visual functional connectivity has been consistently associated with poorer measures of executive function (Rosenthal et al., 2018). It is therefore unsurprising that this network would be the sole network demonstrating significant hypoconnectivity, since it is likely that these critical functions are disturbed following injury. Nonetheless, future studies should include assessment related to the corresponding networks we wish to study, such as visual tests, in order to more accurately determine the extent of impairment following mTBI.

The sensorimotor network, which was one of our IFCNs demonstrating increased functional connectivity, plays an important role in disease-related functional changes in task performance (D’Souza et al., 2020). Enhanced functional connectivity in this network has been correlated with longer recovery time, which may serve as a useful marker for athletes looking to return to sport (Churchill et al., 2017). Increased functional connectivity was also found between the dorsal attention network and the cingulate gyrus, part of the DMN.

In line with our findings, multiple studies (Nathan et al., 2015; Rigon et al., 2016; Sharp et al., 2011; Stevens et al., 2012) have shown that their mTBI group exhibited increased connectivity in the DMN, specifically in the precuneus. Several reports have outlined the vital role of the precuneus in assisting in various behavioural tasks, such as autobiographical memory retrieval, manipulation of mental images, internally guided attention, and reward outcome monitoring (Smitha et al., 2017). Enhanced functional connectivity in the DMN has been elucidated as the brain’s response to inadequately mediate behaviour, due to limitations imposed by the hypothesized impairments to typical neuronal function that accompany mTBI (Sharp et al., 2011; Stevens et al., 2012). It has been suggested in numerous studies that this is precisely what causes the entire network to “compensate”, and this process may begin as early as 10 days post-injury (Palacios et al. 2017; Rosenthal et al., 2018; Stevens et al. 2012). The greater DMN functional connectivity at rest could promote appropriate deactivations of other networks, necessary for focused, goal-directed behaviour (Sharp et al., 2011). It has also been postulated that posterior DMN activity may be part of normal recovery, since the existing literature has found the opposite to be associated with persistent post-traumatic complaints (Orr et al., 2016).

It is worth mentioning that not all of our findings were consistent with previous literature that used ICA analysis to examine the effects of mTBI. Palacios and coworkers (2017) found that DMN and dorsal attentional network were among the networks that showed reductions in connectivity, while our results showed increased connectivity in these IFCNs. This discrepancy might be explained by the window between injury and scan, which in this specific study, was 3-18 days post-injury. Furthermore, the increased functional connectivity in the dorsal attentional network was found when comparing CT/MRI positive mTBI patients to controls, whereas our study had a strict exclusion criterion regarding the presence of structural abnormalities on CT/MRI. Additionally, our results of increased functional connectivity in the sensorimotor network contrasts with the results reported by D’Souza et al. (2020) who found significantly decreased functional connectivity in this region, along with the DMN. However, this reduced connectivity was observed within seven days of sustaining mTBI, and a significant improvement in connectivity was found when assessed six months post injury. This reiterates the notion that timing of imaging after injury remains an important factor to explain variability across studies.

Both ICA and seed-to-voxel analyses are widely used methods in rs-fMRI, but may present conflicting findings, which was the case in our study. When using the spatial match-to-template approach, we found no significant group differences for the SN, the frontoparietal network, or the cerebellar network, despite our findings for the SN in the seed-to-voxel analysis. This raises caution for the interpretation of our voxel-based findings. What is even more unusual is that opposing functional connectivity group differences were found in the DMN, when comparing the ICA results to the seed-to-voxel results. As it may be expected, the differences between these sets of findings are likely a function of different analytic approaches and enforces why we chose to include multiple methods of analysis. It is also important to consider that in the seed-to-voxel analysis, significant findings were extracted based on predefined seeds within a network, whereas ICA analysis considers the network as a single entity, which may also influence the direction of functional connectivity changes.

### 4.4 Correlation with post-concussive symptoms

In terms of correlation with post-concussive symptoms, we found a significant negative relationship between the dorsal attentional network and behavioural symptoms score. It is worth noting that a significant correlation was only observed when taking age and sex as confounds, which emphasizes the importance of this consideration. Because depression was one of the four behavioural symptoms assessed, it is possible that the dorsal attentional network was the one most significantly affected by depression. In fact, in a meta-analysis assessing large scale network dysfunction using rs-fMRI, individuals with major depressive disorder were found to have hypoconnectivity within the dorsal attentional network seeds (Kaiser et al., 2015). Although correlational analysis was run by assessing the network as a whole rather than individual seed points within networks, the nodes automatically included in the CONN toolbox for the dorsal attentional network are the intraparietal sulcus and the frontal eye fields. These two areas are active when attention is overtly or covertly oriented in space (Vossel et al., 2014); this may help explain why there was a correlation with behavioural symptoms score, as the score also includes symptoms such as irritability, frustration, and restlessness. It is therefore possible that these very emotions cause us to orient ourselves towards our external environment, explaining why the correlation was observed.

### 4.5 Hyperconnectivity

Overall, increases in functional connectivity found in our mTBI group across analyses could represent a variety of types of neural compensation or protective mechanisms following injury. However, whether they are directly related to damaged neurons, or concomitant to increased awareness of the external environment, psychological distress, pain or other recovery-related factors remains unclear (Stevens et al., 2012). Secondary responses such as neuroinflammation, edema, disruption of the blood brain barrier, reduced cerebral blood flood could also contribute to delayed effects on functional connectivity (Kaushal et al., 2019). It would be interesting to investigate whether recruitment of additional neural resources following network disruption is only needed at specific time points following recovery; some have associated hyperconnectivity with less severe injury, while others associate it with prolonged symptoms (Kaushal et al., 2019). The nature of increased functional connectivity is likely dependent on several factors such as age, pre-injury cognitive function or availability of neural resources (Stern, 2002). We are therefore unable to establish a link between our observations and a precise pathophysiology. However, future longitudinal work, combined with other neuroimaging techniques, can help pinpoint the neurological and metabolic consequences of mTBI and their effects on functional connectivity.

### 4.6 Limitations

This study further broadens the knowledge and information available from previous literature in several ways, through its examination of numerous networks, the association of functional connectivity with specific categories of post-concussive symptoms, and the use of three analytical methods. Nevertheless, it important to consider the limitations of our findings. First, all patients in our analysis were grouped, even though different symptoms may lead to discrepancies in regional brain damage; thus, it is possible that our findings only reflect common symptoms that are prevalent across varying mTBI types. Although we dealt with many impending influences, factors such as hormones, medication use, aging and stress may also inevitability play a role in variability between patients. Future studies should rigorously report any pharmacological therapy use and investigate the role that these may play on post-concussive symptoms. We also cannot exclude the possibility that establishment of symptom severity through self-reporting introduces bias (Wu et al., 2016). Next, even though disruptions of brain function can emerge overtime and manifest months later, some suggest that three months following injury may be an early time point for significant improvement to occur in connectivity (D’Souza et al. 2020). Therefore, follow-up scans need to be administered to better understand the physiological basis of recovery over time. Lastly, despite our incorporation of multiple networks, there exists a vast number of others that have not been considered in our analysis, that may nonetheless contribute to the underlying pathophysiology of mTBI. Thus, analysis methods that include additional and less recurrent networks may prove useful.

## 5. Conclusion

The overall findings of our study show extensive changes in functional connectivity between and within IFCNs and other brain regions following mTBI, with a less prevalent but nonetheless important correlation with behavioural symptoms. These results highlight that rs-fMRI in mTBI patients may be deemed useful for objectively assessing disrupted functional connectivity among IFCNs that are otherwise difficult to explain with conventional imaging. This technique may also be potentially useful as an injury-based biomarker in mTBI, especially for those still experiencing post-concussive symptoms, long after the acute setting, as well as for athletes looking to safely return to play. With access to rs-fMRI, the clinician will eventually be able to modify its use from a primarily research-focused modality to a more routine clinical tool, with the aim to improve diagnosis, prognosis, and overall management of mTBI.

## Data Availability

Data will be made openly available from the corresponding author upon reasonable request.

## Acknowledgements

We would like to thank Dr. Chandan Kakkar and Dr. Shiasta Riaz, Fellows in Radiology at McGill University, who helped with patient recruitment. We would also like to thank the Fonds de recherche du Québec – Santé (33299) and the McGill Neurosurgery Practice Plan for providing funding that helped to carry out this study. The role of the sponsors had no further involvement in the study. None of the authors have any conflicts of interest to declare.

